# Systematic Identification of Core Targets ABCB1, PIM2, and TSHR Mediating Bisphenol S-Promoted Cutaneous Melanoma Metastasis and Prognostic Model Construction

**DOI:** 10.64898/2026.07.11.26357805

**Authors:** Yuzhen Xiong, Yanting Yu, Chong Zhao

## Abstract

**Background:** Cutaneous melanoma is the most aggressive malignant skin tumor, and metastasis represents the primary cause of patient mortality. Bisphenol S (BPS) has an unclear influence on melanoma metastasis and its underlying molecular mechanisms.

**Methods:** Potential BPS targets were predicted using the SEA, SwissTargetPrediction, and SuperPred databases. Based on TCGA-SKCM transcriptomic data, differential expression analysis was performed, and Weighted Gene Co-expression Network Analysis (WGCNA) was employed to construct a gene co-expression network. Candidate genes were obtained by integrating BPS-related targets, differentially expressed genes (DEGs), module genes, and univariate Cox regression genes, followed by Gene Ontology (GO)/Kyoto Encyclopedia of Genes and Genomes (KEGG) enrichment analysis and protein–protein interaction (PPI) network construction. Least Absolute Shrinkage and Selection Operator (LASSO)-Cox regression was applied to screen core prognostic genes and construct a risk prediction model. Further analyses included network construction, molecular docking, and 100 ns molecular dynamics (MD) simulation.

**Results:** Integration of BPS-related targets, DEGs, WGCNA module genes, and Cox regression results yielded 13 candidate genes enriched in kinase activity regulation and melanoma-related pathways. LASSO-Cox regression ultimately identified three core prognostic genes—ABCB1, PIM2, and TSHR—all significantly upregulated in metastatic tissues, with area under the curve (AUC) values of approximately 0.7. High-expression patients exhibited significantly better overall survival than low-expression patients (P < 0.05). A nomogram incorporating the three genes and clinical parameters demonstrated good calibration performance. Within the ceRNA network, MALAT1 and hsa-miR-155-5p were identified as key regulatory molecules, and 37 potential transcription factors were predicted, including CEBPA, JUN, and STAT3. Molecular docking revealed strong binding affinities of BPS toward ABCB1, PIM2, and TSHR, and MD simulations confirmed the structural stability of all three complexes.

**Conclusion:** ABCB1, PIM2, and TSHR are the core target genes through which BPS influences melanoma metastasis via multidrug resistance, kinase signaling, and receptor-mediated signal transduction. The prognostic model based on these three genes demonstrates good clinical applicability, and the ceRNA and transcription factor regulatory networks provide a systematic molecular basis for understanding the association between BPS exposure and melanoma metastasis.

## 1 Introduction

Cutaneous melanoma, arising from the malignant transformation of epidermal melanocytes, ranks among the most aggressive forms of skin cancer ^[1, 2]^. Figures released in GLOBOCAN 2020 put the annual burden at more than 320,000 new diagnoses and upwards of 57,000 deaths worldwide, with disease incidence disproportionately concentrated in Caucasian populations and in geographic regions subject to heavy ultraviolet exposure ^[3]^. When detected early, the disease carries a relatively benign outlook: surgical resection alone yields 5-year survival rates above 98%. The clinical picture changes dramatically once the tumor disseminates, and patients presenting with stage IV metastatic disease face 5-year survival rates that fall below 25% ^[4]^. While the advent of targeted agents against BRAF/MEK and immune checkpoint blockers directed at PD-1/PD-L1 has reshaped treatment outcomes in recent years, both primary unresponsiveness and acquired resistance continue to undermine their long-term efficacy, leaving many patients to eventually progress on therapy ^[5]^. Against this backdrop, dissecting the molecular underpinnings of melanoma dissemination, uncovering new prognostic markers, and pinpointing actionable therapeutic targets have become pressing priorities in melanoma research. Bisphenol S (BPS) has emerged as the dominant industrial replacement for bisphenol A (BPA), and its thermal stability together with antioxidant properties have secured its widespread incorporation into food packaging, thermal receipt paper, personal care formulations, and various industrial materials ^[6]^. With regulatory restrictions on BPA tightening across much of the world, the manufacture and consumption of BPS have expanded rapidly, driving up human exposure on a population-wide scale. BPS possesses endocrine-disrupting activity and can either mimic or oppose the actions of estrogens, androgens, and thyroid hormones, thereby perturbing physiological endocrine signaling ^[7]^. Evidence accumulated from epidemiological surveys and toxicological experiments links BPS exposure to a spectrum of adverse health outcomes, encompassing immune dysfunction, neurodevelopmental impairment, and metabolic disturbance ^[8, 9]^. Several lines of investigation further suggest that BPS may facilitate breast and prostate tumorigenesis by triggering estrogen receptor–mediated pathways, driving cellular proliferation, and suppressing programmed cell death ^[10–12]^. In contrast, whether and how BPS contributes to the metastatic behavior of cutaneous melanoma remains largely uncharted territory, and a systematic mechanistic inquiry is overdue.

Melanoma metastasis unfolds as a multistep biological cascade governed by interconnected processes, including epithelial-mesenchymal transition (EMT), neovascularization, escape from immune surveillance, and metabolic rewiring ^[13]^. By impersonating endogenous hormonal cues, environmental endocrine disruptors have the potential to derail these molecular programs and alter the proliferative, migratory, and invasive properties of tumor cells—though the precise molecular choreography behind such effects is still being worked out ^[14]^.

Motivated by these gaps, the present work integrates network toxicology with large-scale transcriptomic profiling to dissect the molecular circuitry through which BPS exposure may shape the metastatic progression of cutaneous melanoma. Candidate BPS targets were first assembled by cross-referencing predictions from the SEA, SwissTargetPrediction, and SuperPred databases, after which differential expression profiling and WGCNA co-expression network analysis were applied to transcriptomic data drawn from the TCGA-SKCM cohort. We then built a LASSO-Cox risk prediction model to distill a panel of core prognostic genes, whose clinical relevance was examined through expression validation, correlation with clinicopathological parameters, and survival curve analysis. At the mechanistic level, ceRNA network construction and transcription factor prediction were used to map the multi-layered regulatory architecture surrounding these genes, and the direct physical interaction between BPS and its core protein targets was probed through molecular docking coupled with molecular dynamics simulations. Collectively, this work aims to provide a coherent theoretical framework for understanding how BPS exposure intersects with the molecular machinery driving melanoma progression and metastasis.

## 2 Materials and Methods

### 2.1 Data Acquisition

BPS-associated proteins were computationally predicted by querying three independent databases—SEA (http://sea.bkslab.org/), SwissTargetPrediction (http://www.swisstargetprediction.ch/), and SuperPred (https://prediction.charite.de/)—using the canonical SMILES (Simplified Molecular Input Line Entry System) notation of BPS retrieved from PubChem (https://pubchem.ncbi.nlm.nih.gov/). Outputs from all three platforms were merged and deduplicated to yield a consolidated target gene set, which was then visualized as a BPS–target interaction network in Cytoscape (v3.9.1).

Drug-likeness and pharmacokinetic properties of BPS were profiled on the SwissADME platform (http://www.swissadme.ch/) by submitting the SMILES string. Key descriptors evaluated included lipophilicity (LIPO), molecular weight (MW), polarity (POLARITY), aqueous solubility (INSOLU), degree of unsaturation (INSATU), and molecular flexibility (FLEX); oral bioavailability potential was characterized using Lipinski’s rule of five and the bioavailability radar.

RNA-seq transcriptomic data from the TCGA-SKCM (The Cancer Genome Atlas–Skin Cutaneous Melanoma) cohort were downloaded via the GDC portal (https://portal.gdc.cancer.gov/), covering both primary tumor and metastatic samples with expression values normalized as FPKM (Fragments Per Kilobase Million). Paired clinical annotations—including patient age, sex, tumor staging (T, N, M stages, and overall Stage), and survival follow-up data—were retrieved for downstream analyses. Quality filtering removed genes with zero expression in more than 50% of samples and discarded outlier samples from the expression matrix.

### 2.2 Univariate Cox Regression Analysis

To screen genes with a preliminary association with overall survival (OS), univariate Cox proportional hazards regression was applied to the candidate genes using the R survival package, with OS time and status as endpoint variables. Hazard ratios (HRs) with 95% confidence intervals (CIs) and two-sided P-values were derived per gene; those reaching P < 0.05 were advanced to subsequent LASSO-Cox feature selection.

### 2.3 Differential Expression Analysis

Differences in gene expression between metastatic and primary tumor tissues in the TCGA-SKCM dataset were examined with the limma package (v3.62.1). Genes satisfying |log₂ fold change| > 1 and an adjusted P-value [false discovery rate (FDR) < 0.05] were designated differentially expressed genes (DEGs). The global distribution of expression changes was represented in a volcano plot with upregulated and downregulated genes annotated separately, and a heatmap was constructed to display expression patterns among the 20 most significantly altered genes in each direction.

### 2.4 WGCNA

The WGCNA package (version 1.72) was used to construct a gene co-expression network and identify gene modules associated with disease phenotypes. Outlier samples were first removed through sample clustering analysis. A soft-thresholding power of β = 3 was selected to construct a scale-free network, ensuring conformity with scale-free topology (fitting index R² > 0.85). Hierarchical clustering was performed based on the topological overlap matrix (TOM) dissimilarity, and the dynamic tree cut algorithm was applied to identify gene modules, with the minimum module size set to 30, deep split set to 3, and maximum module merging distance set to 0.25. The correlation between each module eigengene and SKCM was calculated to identify significantly correlated modules (P < 0.05).

### 2.5 Functional Enrichment and Protein-Protein Interaction of Candidate Gene

Candidate genes were defined as those at the intersection of DEGs, module genes, and BPS-related target genes and cox genes; the overlap was depicted using the VennDiagram package.

Gene Ontology (GO) and Kyoto Encyclopedia of Genes and Genomes (KEGG) enrichment analyses were performed on the candidate set using clusterProfiler (v4.8.3), spanning three GO domains—Biological Process (BP), Cellular Component (CC), and Molecular Function (MF)—with enrichment significance assessed by the hypergeometric distribution test (FDR < 0.05). The ten most enriched GO terms and KEGG pathways were summarized as bar charts.

Protein–protein interaction (PPI) network analysis was conducted through STRING (v12.0, https://string-db.org/) at a minimum interaction confidence of 0.4, with the resulting network imported into Cytoscape (v3.9.1). Node degree and betweenness centrality were calculated to identify functionally central hub nodes.

### 2.6 Prognostic Gene Screening and Core Gene Identification

Least Absolute Shrinkage and Selection Operator (LASSO)-Cox regression was applied to the 13 candidate genes for prognostic feature selection. The optimal regularization parameter λ was determined through ten-fold cross-validation at the minimum cross-validation error, and genes retaining non-zero coefficients were designated core prognostic genes. A LASSO coefficient path plot was generated to illustrate the variable selection trajectory.

A risk score formula was constructed based on LASSO-selected genes: Risk Score = Σ(coefficient_i_ × expression_i_). Patients were divided into high- and low-risk groups according to the median risk score. Expression patterns of core genes across risk groups were displayed alongside survival status in a stratified heatmap. Kaplan–Meier (K–M) survival curves were drawn for each core gene at the median expression cutoff, with log-rank tests used to assess inter-group survival differences; a parallel analysis was conducted on risk-score–stratified patient groups to validate model performance.

### 2.7 Core Gene Expression Validation and Diagnostic Value Assessment

Differential expression of the key genes between metastatic and primary tissues was quantified by the Wilcoxon rank-sum test and presented as box plots. Receiver operating characteristic (ROC) curves were constructed per gene, and area under the curve (AUC) values calculated to evaluate diagnostic discriminability. A two-sided P < 0.05 was applied throughout.

### 2.8 Prognostic Model Construction and Nomogram Development

Core gene expression levels were integrated with clinical parameters (gender, T stage, N stage, M stage, and overall Stage) to construct a multivariate Cox regression prognostic model. Regression coefficients informed the construction of a nomogram for individualized survival probability estimation, with each variable assigned a scoring scale reflecting its prognostic contribution. Internal validation employed Bootstrap resampling (1,000 iterations), and calibration curves at 1, 3, and 5 years were plotted to assess concordance between predicted and observed outcomes.

### 2.9 Competing Endogenous RNA and Transcription Factor Networks Construction

The competing endogenous RNA (ceRNA) regulatory network for the three core genes was assembled via the miRNet database (https://www.mirnet.ca/). miRNA binding partners for each core mRNA were first retrieved, followed by identification of upstream lncRNAs interacting with those miRNAs. The complete network was rendered in Cytoscape (v3.9.1), with node degree used to rank key regulatory hubs. Transcription factors (TFs) regulating the three core genes were sourced from the TRRUST v2 database, which archives literature-validated human TF–target relationships. Enrichment significance of the retrieved TF set was evaluated by hypergeometric testing (P < 0.05), and the regulatory network was visualized in Cytoscape (v3.9.1).

### 2.10 Molecular Docking Analysis and Molecular Dynamics Simulation

The three-dimensional structural file of BPS (SDF format) was downloaded from PubChem. Crystal structures of target proteins were sourced from the Protein Data Bank (PDB, https://www.rcsb.org/); where experimental structures were unavailable, AlphaFold3-predicted or homology-modeled structures were substituted. Pre-docking preparation in AutoDockTools (v1.5.7) encompassed polar hydrogen addition, Gasteiger charge computation, and active site definition. Docking was executed in AutoDock Vina (v1.2.0) at exhaustiveness = 8; the lowest-energy binding pose was selected and rendered three-dimensionally in PyMOL (v2.5), with two-dimensional interaction diagrams (hydrogen bonds, hydrophobic contacts, and π–π stacking) generated in Discovery Studio.

For molecular dynamics (MD) simulations, protein geometries from AlphaFold3 and small-molecule conformations from PubChem were adopted. PM3-level atomic charges were assigned via MOPAC; ligand preparation in AutoDockTools 1.5.6 covered polar hydrogen addition, Gasteiger charge assignment, rotatable bond identification, and pdbqt generation. Re-docking in AutoDock Vina used a protein-centered grid of 100 × 100 × 100 points, 50 docking runs, and a maximum of 20,000 iterations. Top-ranked complexes were energy-minimized under the Amber14 force field (1,000 steepest descent + 5,000 conjugate gradient steps) and inspected in Discovery Studio for key interaction contacts. One-hundred-nanosecond MD simulations were subsequently run in GROMACS 2021 under the Amber14SB force field, with complex stability evaluated by root mean square deviation (RMSD), radius of gyration (Rg), solvent accessible surface area (SASA), and MM-GBSA binding free energy decomposition.

### 2.11 Statistical Analysis

All analyses were carried out in R (v4.3.0). Continuous variables were compared by the Wilcoxon rank-sum test or Student’s t-test as appropriate; categorical variables were analyzed by chi-square or Fisher’s exact test. Survival analyses followed the K–M method with log-rank testing; Cox regression was employed for prognostic factor evaluation. Multiple testing was corrected by the Benjamini–Hochberg (BH) procedure. Correlation analyses applied Pearson or Spearman coefficients depending on data distribution. A two-sided P < 0.05 was considered statistically significant. All figures were rendered using ggplot2 (v3.4.0) and associated visualization packages.

## 3 Results

### 3.1 Preliminary Exploration, Toxicological Characterization, and Impact Analysis of Bisphenol S

The physicochemical properties and oral bioavailability of BPS (C_12_H_10_O_4_S, MW = 250.27 g/mol) were evaluated using the SwissADME platform. SwissADME analysis revealed that BPS exhibits favorable oral bioavailability, adequate aqueous solubility, high gastrointestinal absorption, no CYP450 inhibitory activity, and satisfactory drug-likeness profiles, suggesting efficient systemic absorption potential upon human exposure (Figure 1). These results suggest that BPS possesses a well-balanced pharmacokinetic profile with efficient systemic absorption potential, highlighting the necessity for further toxicological characterization given its widespread environmental prevalence.

### 3.2 Identification of Potential Targets and Pathways Associated with BPS Exposure

By searching and integrating the SEA, SwissTargetPrediction, and SuperPred databases, 145 BPS-related targets (including PIM2, ABCB1, MAOA, and TSHR) were identified (Figure 2).

### 3.3 Identification of Candidate Genes

Univariate Cox proportional hazards regression analysis was performed, and a total of 2,955 cox genes were significantly associated with OS (P < 0.05). Differential expression analysis showed that compared with primary tumor tissues, a total of 9,318 genes exhibited significantly altered expression levels in metastatic tumor tissues, of which 7,401 genes were significantly upregulated and 1,917 genes were significantly downregulated (Figure 3A, 3B).

WGCNA was performed on the transcriptomic dataset to identify gene modules and their constituent genes significantly associated with the disease grouping (Primary Tumor vs. Metastatic). Specifically, the optimal soft-thresholding power β = 3 was selected, at which the scale-free topology fitting index R² reached 0.86 (Figure 4A). Based on hierarchical clustering and the dynamic tree cut algorithm, a total of 10 gene modules were generated (Figure 4B, 4C). Among these, 7 modules were significantly correlated with the SKCM (P < 0.05) (Figure 4D). 7,512 module genes were extracted and identified from these significant gene modules. The intersection of DEGs, module genes, cox genes and BPS-related genes yielded 13 candidate genes (Figure 4E).

### 3.5 Functional Enrichment Analysis and PPI Network

To further investigate the functional roles of the candidate genes in disease pathogenesis, systematic functional enrichment analysis was performed. GO enrichment analysis revealed that BPS target genes were significantly enriched in kinase-related biological processes (positive regulation of kinase activity and protein kinase activity) and molecular functions closely associated with melanoma pathogenesis, including protein tyrosine kinase activity, transmembrane receptor protein tyrosine kinase activity, and protein serine/threonine kinase activity (Figure 5A–C). Notably, pathway enrichment analysis further identified the melanoma signaling pathway as one of the most significantly enriched pathways, alongside the cAMP signaling pathway which plays a critical regulatory role in melanocyte proliferation and differentiation, collectively suggesting that BPS may promote melanoma development and progression through dysregulation of tyrosine kinase-mediated signaling cascades (Figure 5D–E).

To further elucidate the interactions among the candidate genes, a PPI network was constructed (Figure 5F), revealing extensive and close interactions among ABCB1, FLT3, SIRT1 and CDK6, suggesting their coordinated involvement in disease pathological processes.

### 3.6 Key Gene Screening

LASSO-Cox regression applied to the 13 candidate genes demonstrated progressive coefficient shrinkage with increasing λ. Ten-fold cross-validation identified the optimal λ as 0.11 (Figure 6A), at which three genes—ABCB1, PIM2, and TSHR—retained non-zero coefficients and were designated as core prognostic features. Risk-stratified heatmap analysis showed markedly divergent expression profiles between high- and low-risk patients, with a concordant gradient in survival status (Figure 6B). K–M analysis confirmed that patients with high expression of each gene had significantly better OS than those with low expression (all P < 0.05; Figures 6C–E). Model-based risk stratification additionally demonstrated that low-risk patients carried a substantially more favorable prognosis than high-risk counterparts, validating the selected gene panel (Figure 6F).

### 3.7 Key Gene Expression Validation

The Wilcoxon rank-sum test was used to analyze the expression patterns of feature genes ABCB1, PIM2, and TSHR, along with their ROC. All three core genes were significantly upregulated in the metastatic group, with AUC values approximating 0.7 for the ROC, indicating moderate diagnostic capability (Figures 7A, 7B).

To construct a clinically applicable risk prediction tool, ABCB1, PIM2, and TSHR were integrated with clinical information including gender and pathological features (T stage, N stage, M stage, and overall Stage). In this model, both clinical variables and gene expression levels were incorporated as risk factors, with the scoring scale for each variable intuitively reflecting its contribution to prognosis. The total score obtained by summing individual variable scores enables prediction of patient survival probability (Figure 7C). Calibration curve analysis demonstrated good agreement between predicted and actual observed probabilities (Figure 7D), indicating that the nomogram model has high predictive accuracy and is suitable for clinical prognostic evaluation. Notably, although the three genes were upregulated in metastatic lesions, their high expression was associated with more favorable overall survival, a dissociation discussed in Section 4.

### 3.8 Regulatory Network of Core Genes

We investigated the regulatory mechanisms of key genes in the development and progression of melanoma. The ceRNA network comprised 3 key mRNAs, 17 miRNAs, and 34 lncRNAs. Notably, MALAT1 carried the highest degree value; hsa-miR-155-5p ranked highest among miRNA nodes; reciprocal regulatory interactions between these two hub molecules were identified within the network (Figure 8A). This suggests that the ceRNA network may represent a key regulatory mechanism through which BPS influences melanoma metastasis.

TF enrichment analysis was subsequently performed on the key genes ABCB1, PIM2, and TSHR. 37 potential TFs were predicted to regulate at least one of the three key genes based on TRRUST curated relationships, including CEBPA, JUN, and STAT3 (Figure 8B).

### 3.9 Molecular Docking and Molecular Dynamics Simulation

Molecular docking confirmed direct binding of BPS to all three core proteins, with calculated free energies of −7.8 kcal/mol (BPS–ABCB1), −7.4 kcal/mol (BPS–PIM2), and −7.0 kcal/mol (BPS–TSHR) (Supplementary Table S1). In each complex, BPS occupied the binding pocket formed by key amino acid residues, stabilized through a combination of hydrogen bonds and hydrophobic contacts (Figures 9A–C), providing a structural basis for direct BPS-mediated modulation of these targets.

Long-term binding stability was further examined by 100 ns MD simulations in GROMACS 2021 under the Amber14SB force field (Supplementary Table S2). RMSD analysis showed rapid stabilization: ABCB1–BPS and PIM2–BPS complexes converged at approximately 0.2 nm, and TSHR–BPS at approximately 0.4 nm. Rg values remained essentially constant throughout, while SASA exhibited a progressive decline, collectively indicating that all three complexes maintained compact, stable conformations over the full simulation (Figures 10A–C). Together, the docking and MD data provide consistent structural evidence supporting the direct physical engagement of BPS with ABCB1, PIM2, and TSHR.

## 4. Discussion

This study investigated the potential effects of BPS exposure on the melanoma metastasis process by integrating network toxicology, transcriptomics analysis, and machine learning to identify key targets and regulatory mechanisms of BPS intervention in melanoma metastasis. The study identified three core prognostic genes—ABCB1, PIM2, and TSHR—whose high expression was significantly associated with better overall survival, and which demonstrated good calibration in the line graph model. ceRNA network analysis revealed that MALAT1, hsa-miR-155-5p, and transcription factors such as CEBPA, JUN, and STAT3 play key roles in regulating the expression of these core genes. Finally, molecular docking and 100 ns molecular dynamics simulations further demonstrated stable binding between BPS and the three targets.

GO enrichment analysis revealed that BPS-associated candidate genes are enriched in molecular functions such as protein tyrosine kinase activity, transmembrane receptor protein tyrosine kinase activity, and protein serine/threonine kinase activity. Studies have shown that inhibiting the abnormal activation of receptor tyrosine kinases can effectively improve melanoma progression ^[15, 16]^. Furthermore, the activation of serine/threonine kinases also promotes tumor proliferation and metastasis ^[17]^, and targeted therapies against serine/threonine kinases hold potential for treating advanced melanoma ^[18]^. The key target of this study, PIM2, belongs to the serine/threonine kinase family ^[19]^. These kinases maintain tumor cell survival and promote their invasiveness ^[20, 21]^, and the enrichment results are consistent with the functional role of PIM2 described below. KEGG pathway analysis further identified enrichments in the melanoma signaling pathway, the hypoxia-inducible factor (HIF) pathway, and the cAMP signaling pathway. The HIF pathway is persistently activated in the melanoma microenvironment; it creates favorable conditions for tumor metastasis by promoting tumor angiogenesis, inducing the epithelial-mesenchymal transition (EMT) process, and simultaneously shaping an immunosuppressive microenvironment ^[22]^. The cAMP signaling pathway is a key signaling axis regulating the proliferation, differentiation, and survival of normal melanocytes ^[23, 24]^. In this study, the core target TSHR, as a classic Gs-coupled receptor, relies directly on the cAMP-PKA axis for downstream signal transduction ^[25]^, which corroborates the critical role of TSHR in melanoma. ABCB1 (P-glycoprotein) is a core member of the ABC transporter family. It drives substrate efflux through ATP hydrolysis, restricts drug entry into the central nervous system at the blood-brain barrier, and serves as a key molecule mediating classic multidrug resistance in tumors ^[26, 27]^. Overexpression of ABCB1 can directly contribute to resistance to first-line clinical chemotherapy drugs such as doxorubicin and etoposide ^[28]^. Furthermore, its high expression is associated with tumor progression and distant metastasis in patients ^[29]^, which is consistent with the expression validation results of this study. In addition to its role in drug resistance, ABCB1 can also alter cell membrane lipid composition, accelerate the epithelial-mesenchymal transition (EMT) process, and regulate cell migration and invasion ^[30]^. This study found that molecular docking revealed a binding energy of -7.8 kcal/mol between the two, indicating a stable interaction that may synergistically enhance their dual functions of drug resistance and metastasis promotion. However, ABCB1 is also constitutively expressed in peripheral blood lymphocytes and cytotoxic CD8⁺ T cells, where it participates in cytokine transport and lymphocyte function ^[31, 32]^. It can therefore be inferred that the overall elevation of ABCB1 expression in metastatic lesions may partially reflect an increased contribution from infiltrating lymphocytes, which is associated with a relatively favorable prognosis. PIM2 belongs to the PIM family of serine/threonine kinases and participates in downstream regulation of the JAK/STAT pathway, playing a central role in inhibiting apoptosis and promoting protein synthesis ^[33, 34]^. PIM2 can activate the mTOR pathway to mediate lipid metabolic reprogramming, meeting the high energy demands of tumor cells and ensuring their survival and proliferation ^[35]^. In various malignant tumors, PIM2 has been identified as an oncogene, and its high expression is associated with tumor progression, treatment resistance, and poor patient prognosis ^[36]^. BPS stably binds to PIM2, further enhancing kinase activity and improving tumor cell survival and metastatic capacity.

The thyroid-stimulating hormone receptor (TSHR) belongs to the glycoprotein hormone receptor subfamily and is a member of the G protein-coupled receptor (GPCR) superfamily ^[37]^. In thyroid follicular cells, the cAMP pathway mediated by TSHR serves as the central mechanism for the physiological regulation of hormone synthesis, cell proliferation, and differentiation ^[38]^. In melanoma, TSHR may promote cell proliferation and invasion by activating the downstream cAMP/PKA signaling pathway. Concurrently, TSHR expression can be regarded as an alternative marker of the retention of melanocyte lineage characteristics. The progression of melanoma is closely associated with the gradual loss of melanocyte differentiation, which in turn is linked to treatment resistance and poor clinical prognosis ^[39]^. Therefore, the findings of this study suggest that high TSHR expression may indicate that the tumor retains a more differentiated melanocyte phenotype, thereby conferring a better prognosis even in metastatic lesions. Furthermore, TSHR is a key target of environmental endocrine disruptors, and BPS can interfere with normal TSHR signaling by mimicking thyroid hormone ligands. Molecular docking and molecular dynamics simulations in this study confirmed the structural stability of the BPS-TSHR complex. This suggests that BPS may accelerate melanoma progression by continuously activating downstream pro-metastatic signaling through its binding to TSHR. Taken together, the upregulation of ABCB1, PIM2, and TSHR in metastatic lesions, driven by the immune microenvironment, cellular composition, and melanocyte differentiation, is consistent with their favorable prognostic value. However, we acknowledge that future studies involving functional interference experiments are needed to validate the expression patterns and confirm the causal direction of the observed associations.

The ceRNA network constructed in this study encompasses three core mRNAs (ABCB1, PIM2, and TSHR), 17 miRNAs, and 34 lncRNAs, with clearly defined functional roles for the core regulatory nodes. MALAT1, a classic pro-tumor metastasis lncRNA, is highly expressed in various solid tumors and exerts its oncogenic function by competitively binding to miRNAs ^[40–42]^. Network analysis suggests that MALAT1 binds to hsa-miR-155-5p, blocking the miRNA’s interaction with the 3′ UTR regions of its downstream targets ABCB1, PIM2, and TSHR. This releases the miRNA’s repressive effect on core target gene expression, forming a MALAT1/hsa-miR-155-5p/core mRNA regulatory axis. hsa-miR-155-5p is a tumor-suppressing miRNA that specifically inhibits the expression of core pro-metastatic genes while also participating in the regulation of the immune microenvironment ^[43]^. MALAT1’s binding to hsa-miR-155-5p indirectly amplifies the pro-metastatic effects of these core genes, providing a potential target for future RNA-targeted therapies. At the transcriptional regulation level, CEBPA, JUN, and STAT3 are key regulatory molecules. CEBPA is involved in tumor cell differentiation and metabolic regulation ^[44]^ and can bind to the promoter regions of ABCB1 and TSHR to regulate their transcription; its expression is directly correlated with the differentiation grade of melanoma and patient prognosis. As a core member of the AP-1 complex, JUN can directly activate transcription via AP-1 binding sites on the ABCB1 promoter ^[45]^. Persistent activation of the AP-1 pathway has been shown to be clearly associated with tumor drug resistance and invasive progression ^[46]^. STAT3 is a core transcription factor that is persistently activated in tumors and is associated with tumorigenesis, metastasis, and recurrence in various cancers ^[47]^. The regulatory network predicted in this study indicates that it can directly regulate PIM2 transcription and may be involved in mediating melanoma cell anti-apoptosis and metastasis.

In summary, as the primary industrial substitute for bisphenol A, BPS is currently widely present in everyday consumer products such as food contact materials and thermal paper, and exposure levels in the general population are widespread. This study suggests that exposure to BPS at environmentally relevant concentrations may interfere with melanoma cell resistance and metastasis pathways by targeting ABCB1, PIM2, and TSHR, providing preliminary theoretical support for the “environmental exposure–molecular targets–tumor progression” model. This contributes to the further protection of environmental public health and the promotion of clinical translation. This study relies primarily on bioinformatics and computational modeling; all key conclusions require direct experimental validation. The actual binding affinity between BPS and its targets must be confirmed through biochemical methods such as SPR or ITC; the regulatory relationship between the ceRNA axis and transcription factors must be validated via dual-luciferase reporter assays and ChIP-seq; The study data are derived from a single TCGA-SKCM cohort; validation in an independent external cohort is still lacking, and the generalizability of the prognostic model requires cautious evaluation; the actual biological effects at real-world BPS exposure levels also need to be further clarified through epidemiological and toxicological studies. Future work will systematically validate the roles of ABCB1, PIM2, and TSHR in melanoma metastasis at the cellular and animal levels; assess the actual impact of BPS exposure on tumor malignant phenotypes; explore the feasibility of combination therapy with core target inhibitors and immune checkpoint inhibitors; and advance the clinical translation of the prognostic model through prospective cohort studies.

## Author Contributions

Y.X. conceived and designed the study, performed the key bioinformatics analyses and molecular docking experiments, interpreted the main findings, validated the results, provided critical feedback, supervised the study, and drafted and revised the manuscript. Y.Y. collected and processed the data, and contributed to drafting the manuscript. C.Z. assisted with bioinformatics analyses and results interpretation. All authors discussed the results and approved the final manuscript.

## Data Availability

All data used in this study are publicly available. RNA-seq transcriptome data for skin cutaneous melanoma (SKCM) were downloaded from The Cancer Genome Atlas (TCGA) database (https://portal.gdc.cancer.gov/). BPS-related target genes were predicted by integrating results from the SEA database (http://sea.bkslab.org/), SwissTargetPrediction database (http://www.swisstargetprediction.ch/), and SuperPred database (https://prediction.charite.de/). The SMILES notation and three-dimensional structural files of BPS were obtained from the PubChem database (https://pubchem.ncbi.nlm.nih.gov/). Pharmacokinetic and drug-likeness properties of BPS were evaluated using the SwissADME database (http://www.swissadme.ch/). Gene set annotations for functional enrichment analysis were obtained from the Molecular Signatures Database (MSigDB) (https://www.gsea-msigdb.org/). Protein structures of core target proteins were retrieved from the Protein Data Bank (PDB) database (https://www.rcsb.org/). Protein-protein interaction data were obtained from the STRING database v12.0 (https://string-db.org/). Transcription factor regulatory relationships were obtained from the TRRUST database (https://www.grnpedia.org/trrust/), and ceRNA network interactions were predicted using the miRNet database (https://www.mirnet.ca/).

https://sea.bkslab.org/

https://www.swisstargetprediction.ch/

https://prediction.charite.de/

https://pubchem.ncbi.nlm.nih.gov/

https://www.swissadme.ch/

https://www.gsea-msigdb.org/

https://www.rcsb.org/

https://string-db.org/

https://www.grnpedia.org/trrust/

## Acknowledgments

We would like to express our sincere gratitude to all the participants for their contributions to this study.

## Conflicts of Interest

The authors declare no conflicts of interest related to this study.

## Funding

The authors received no financial support for the research, authorship, and/or publication of this article.

Clinical trial number: not applicable.

Ethics, Consent to Participate, and Consent to Publish declarations: not applicable.

---

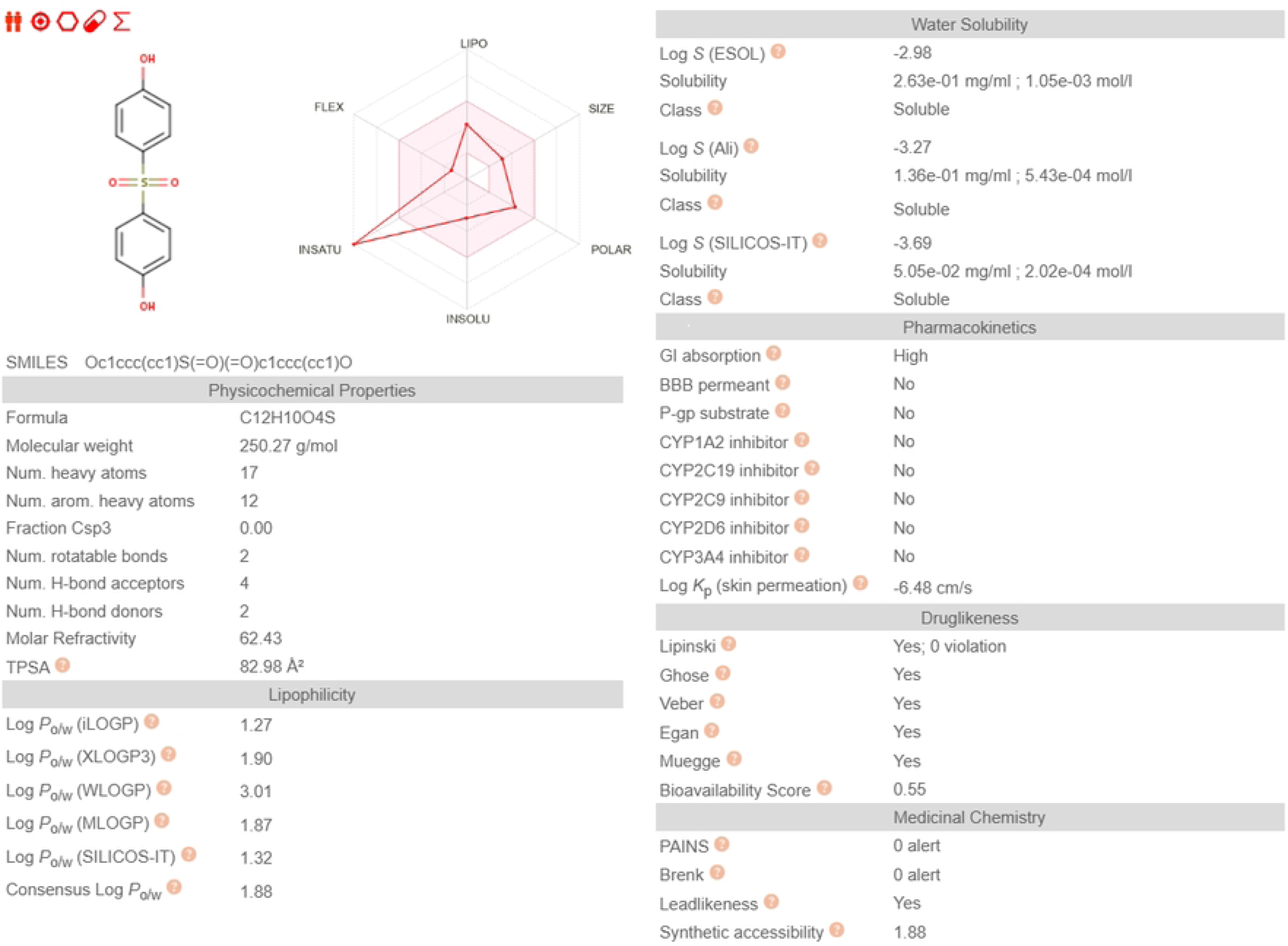

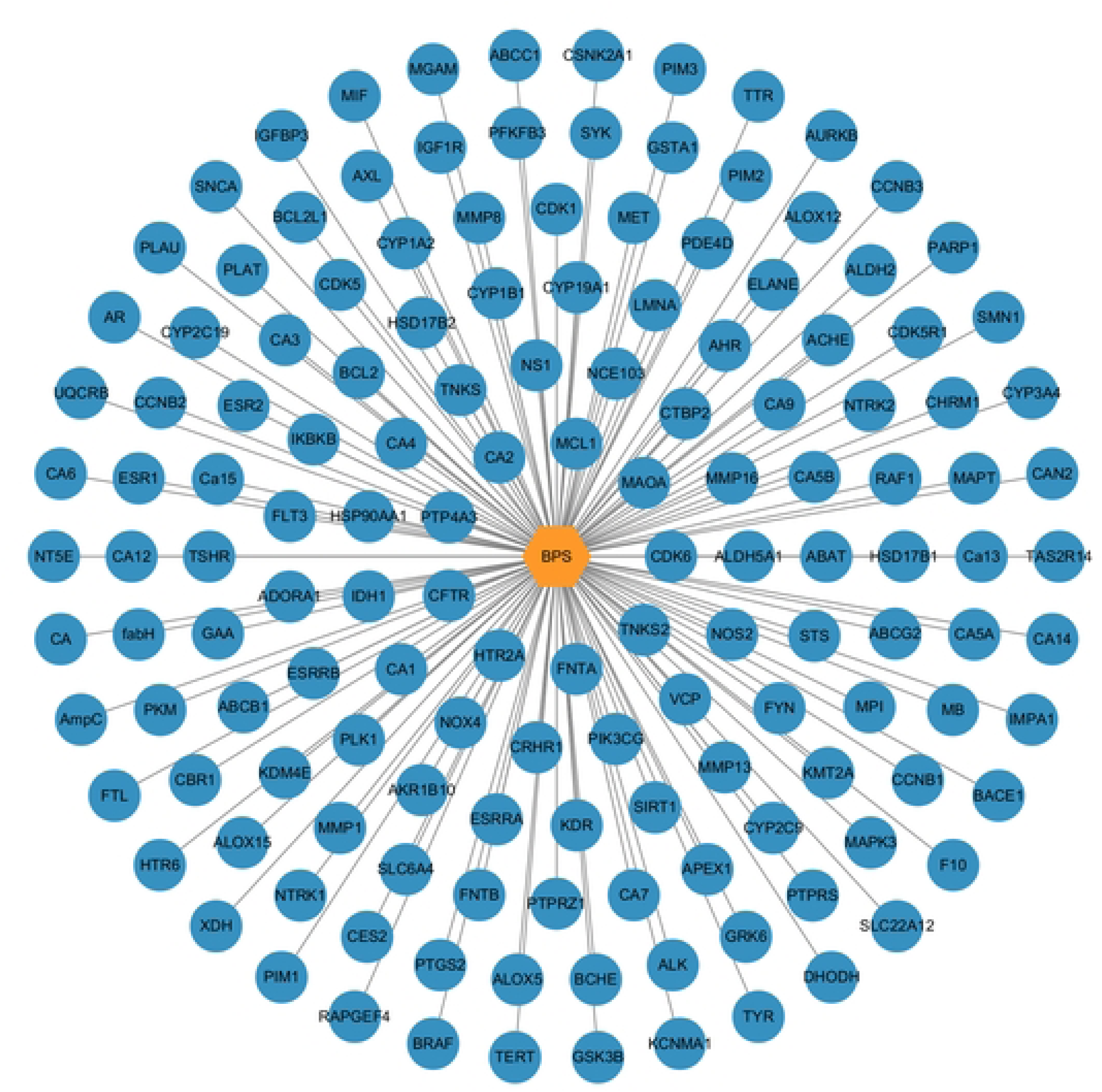

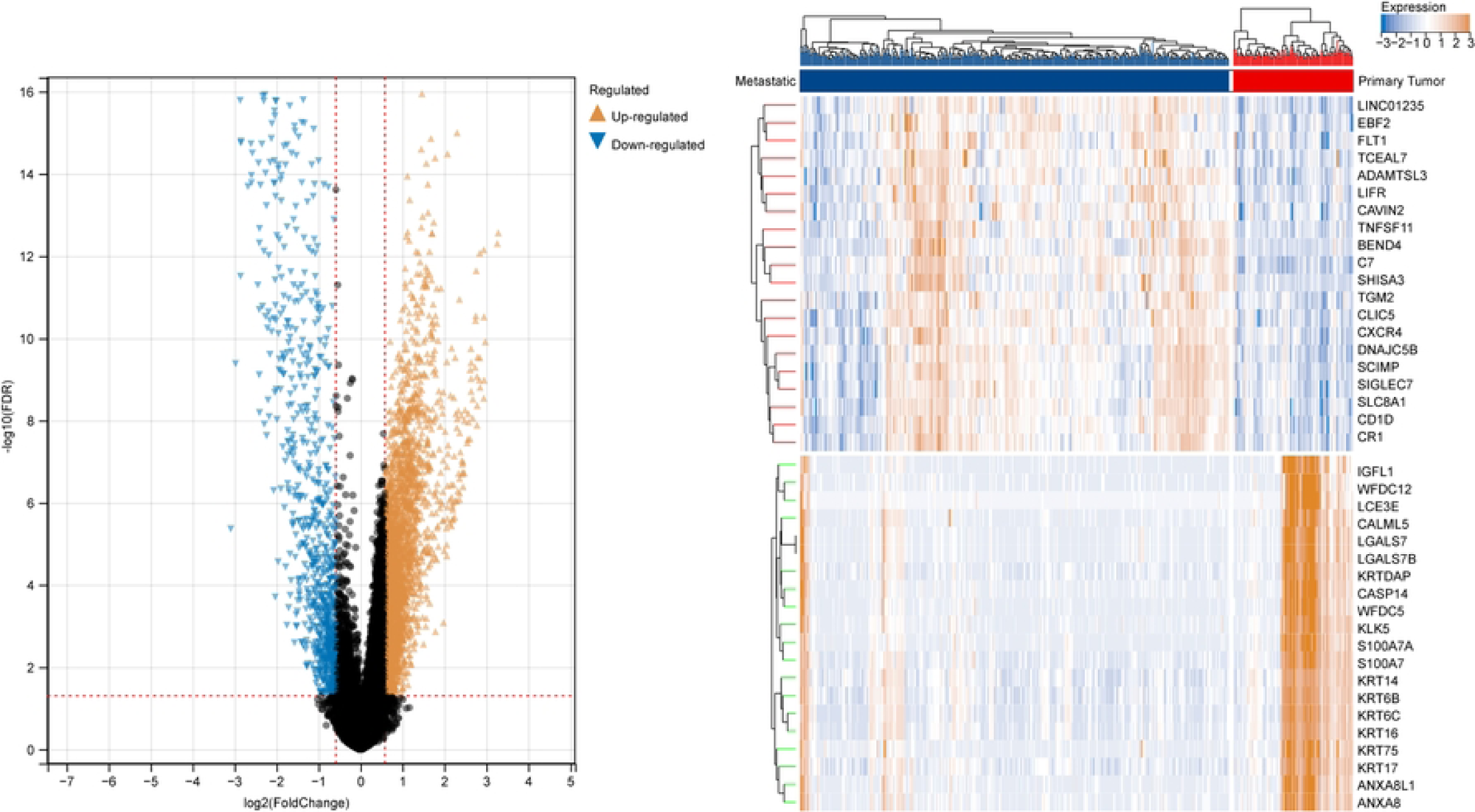

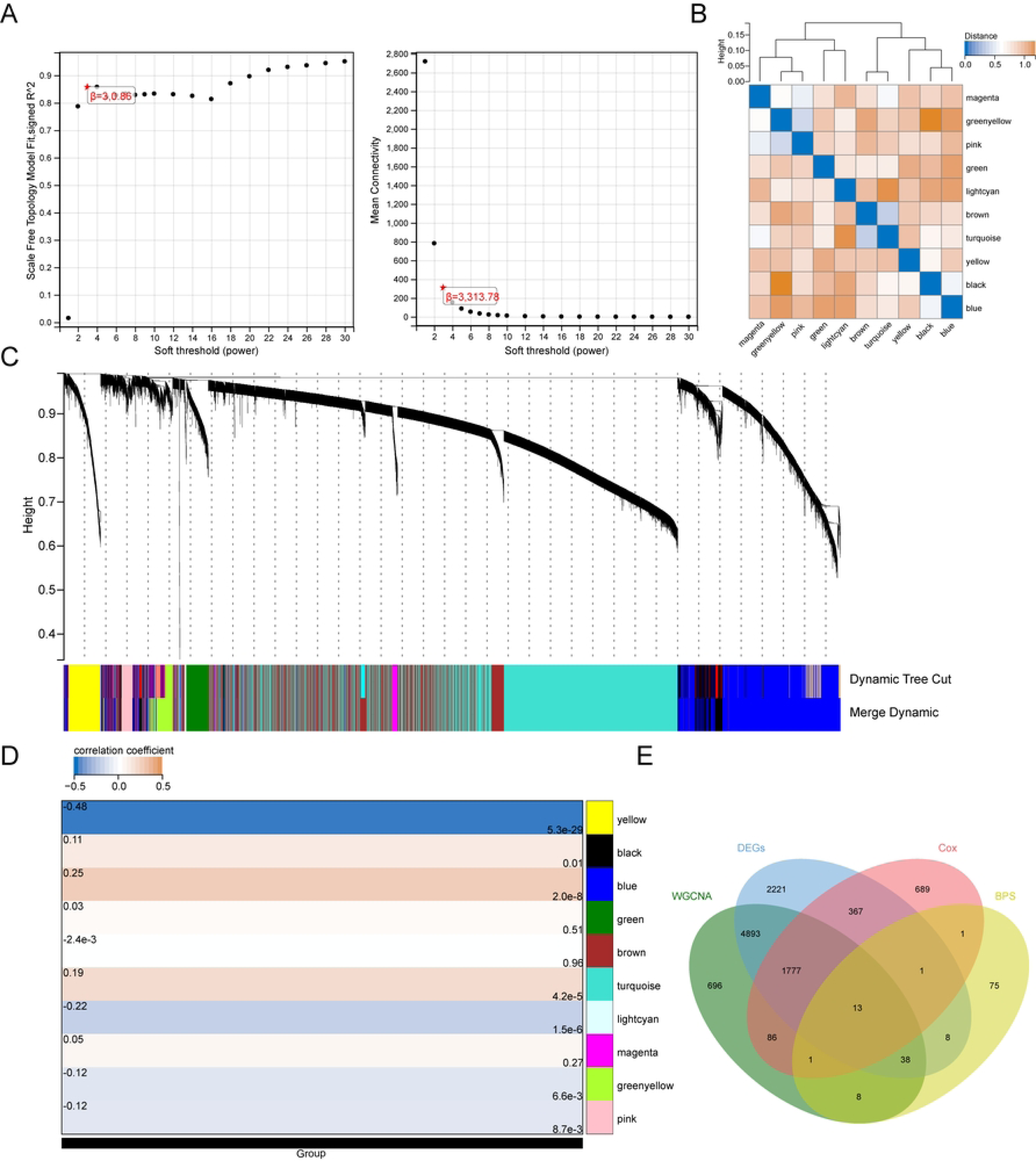

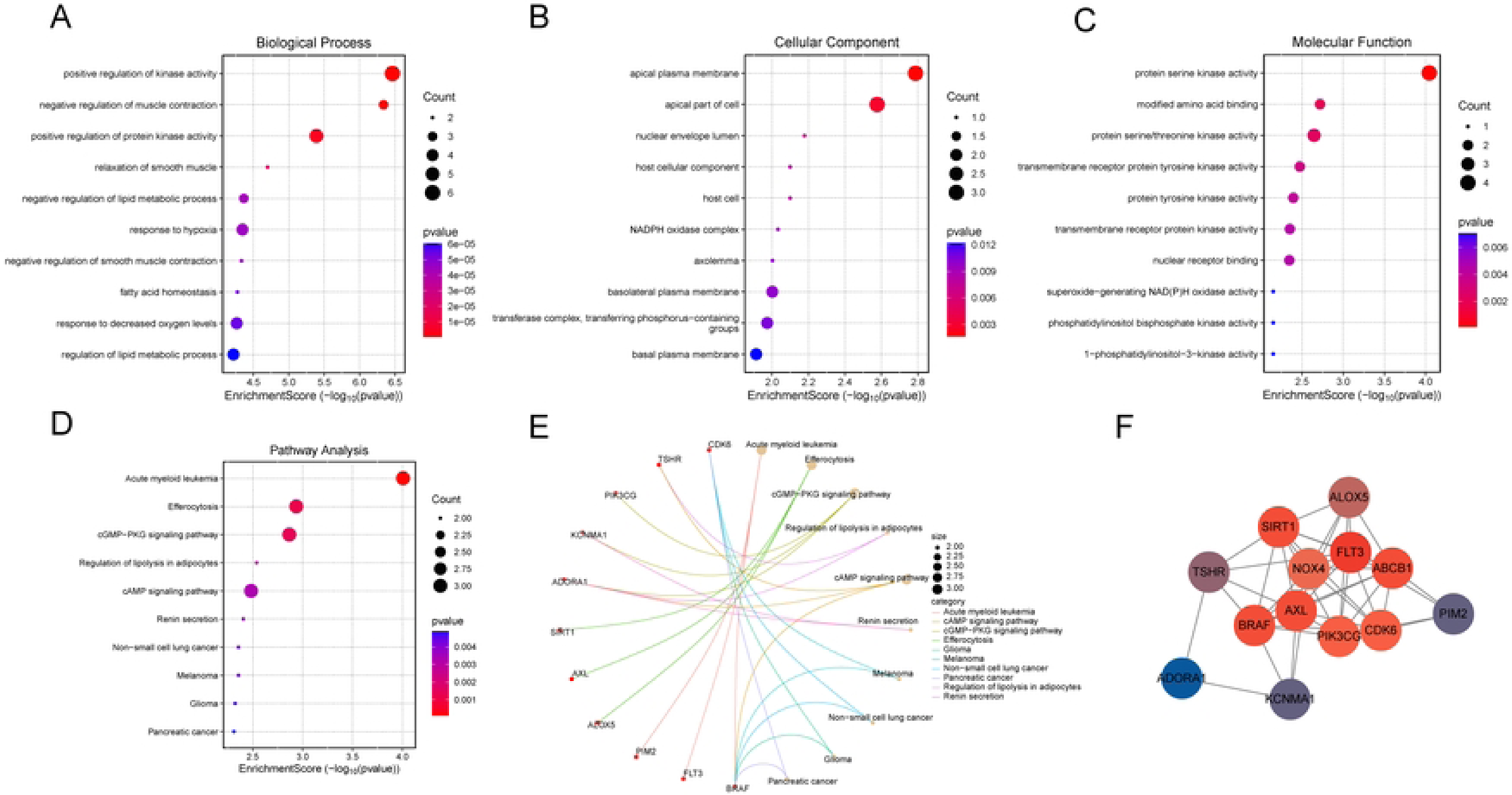

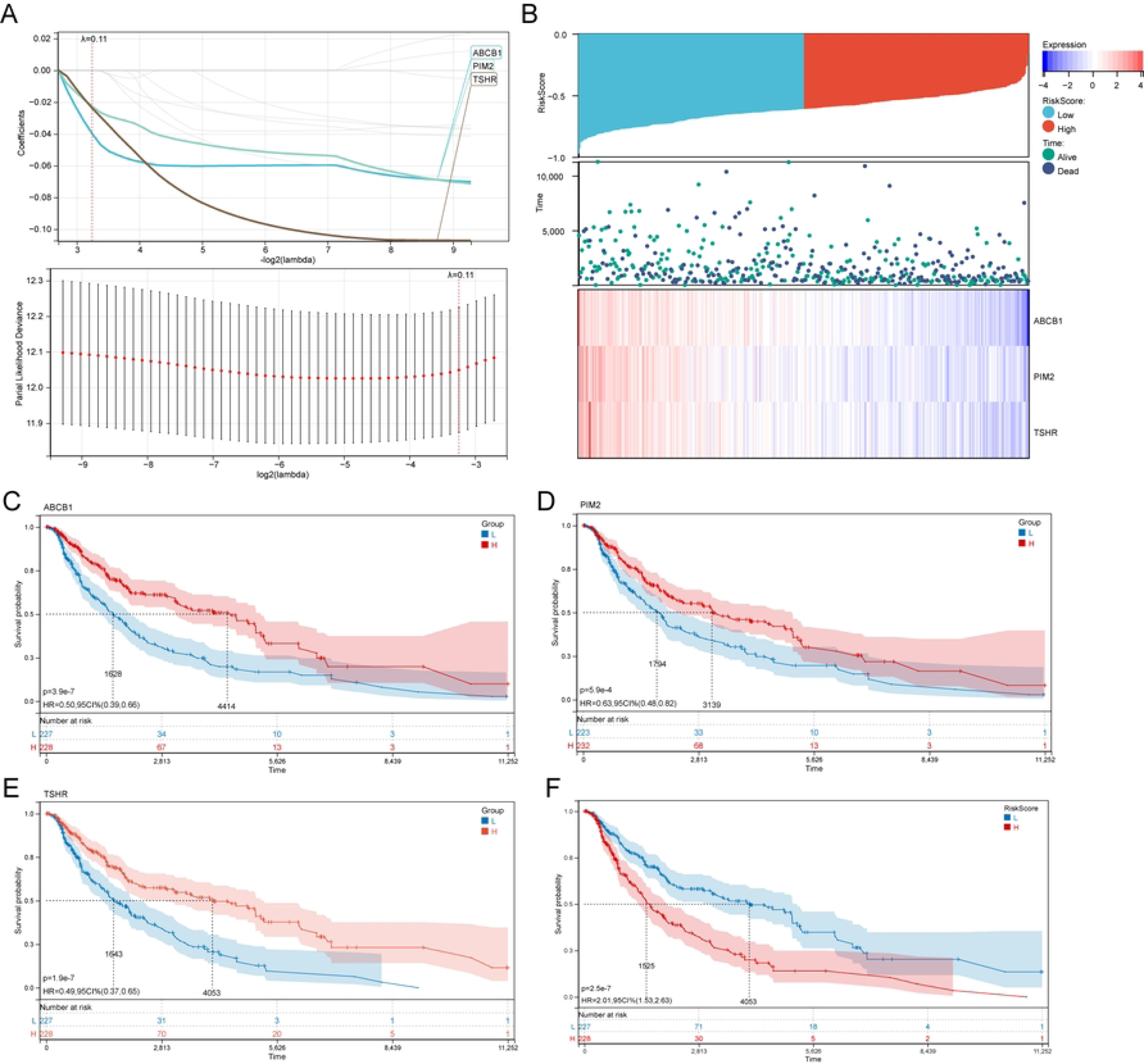

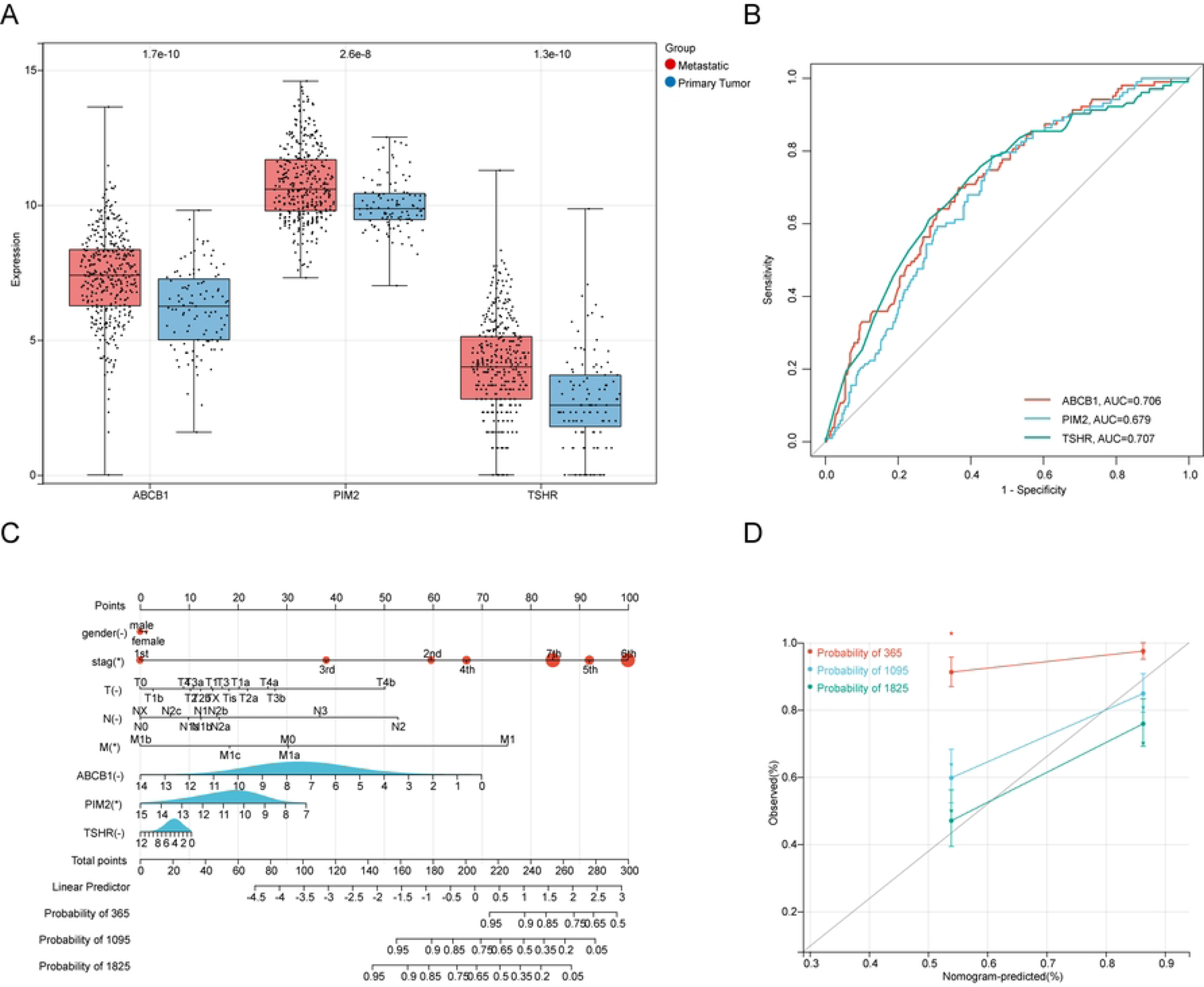

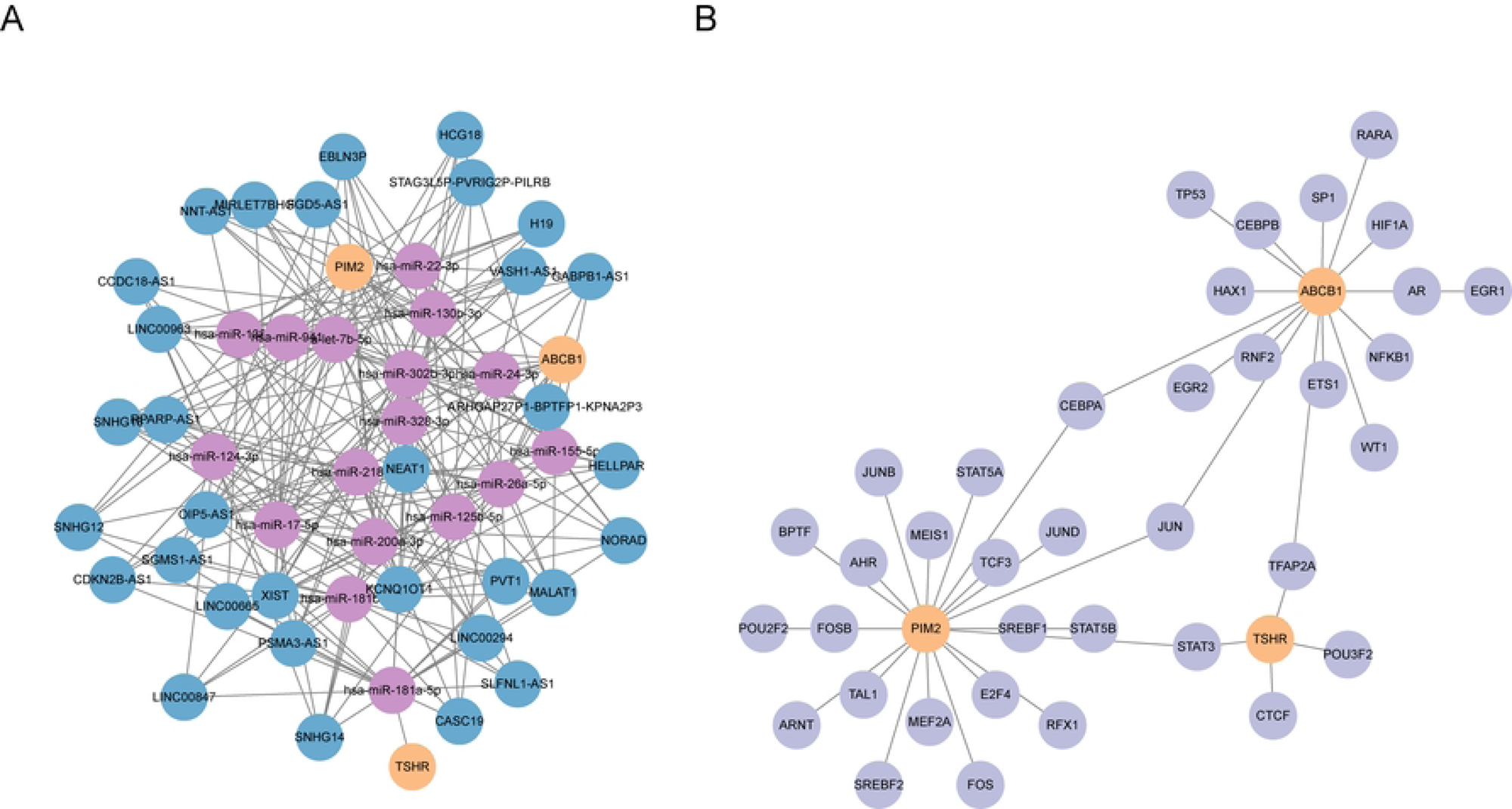

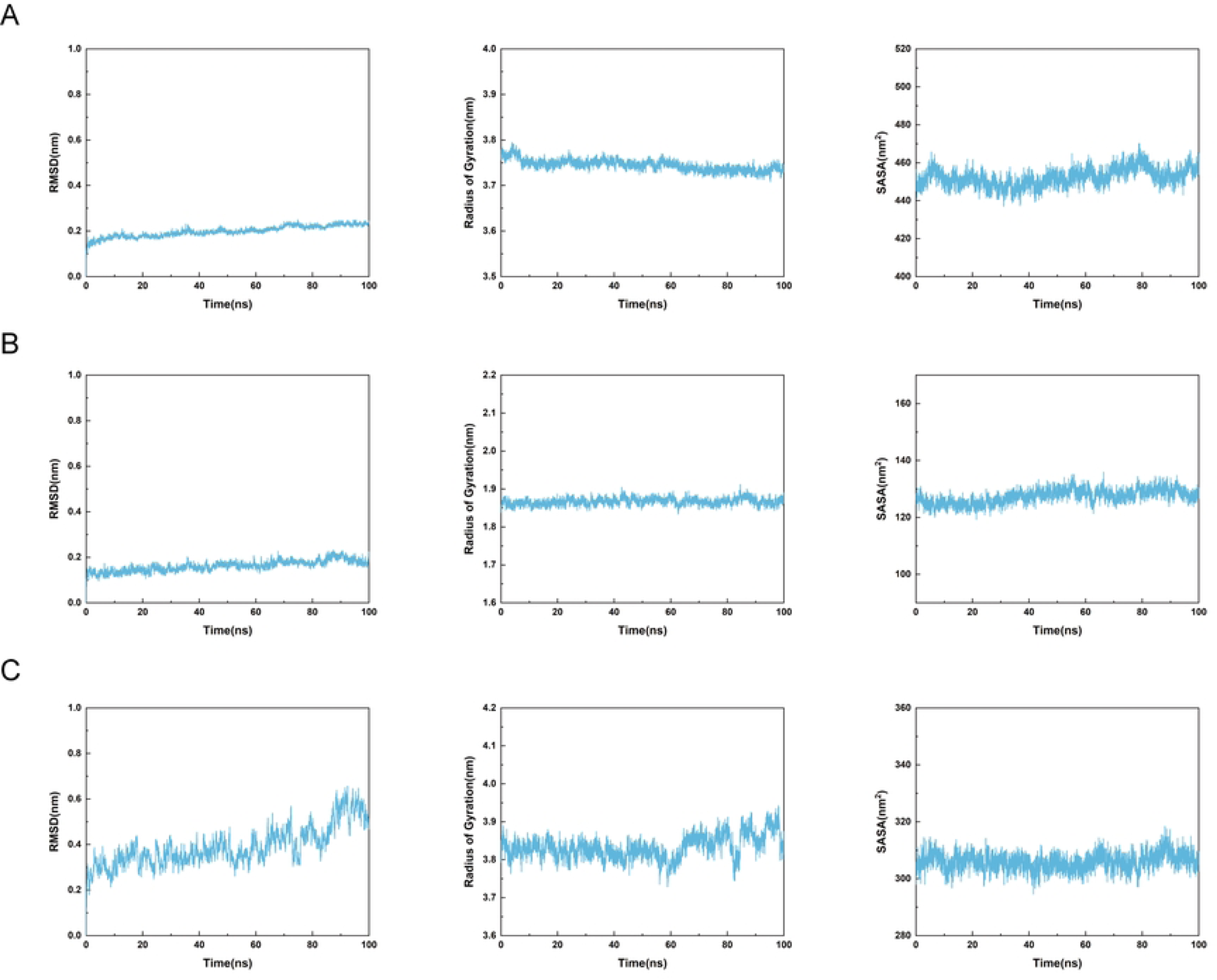

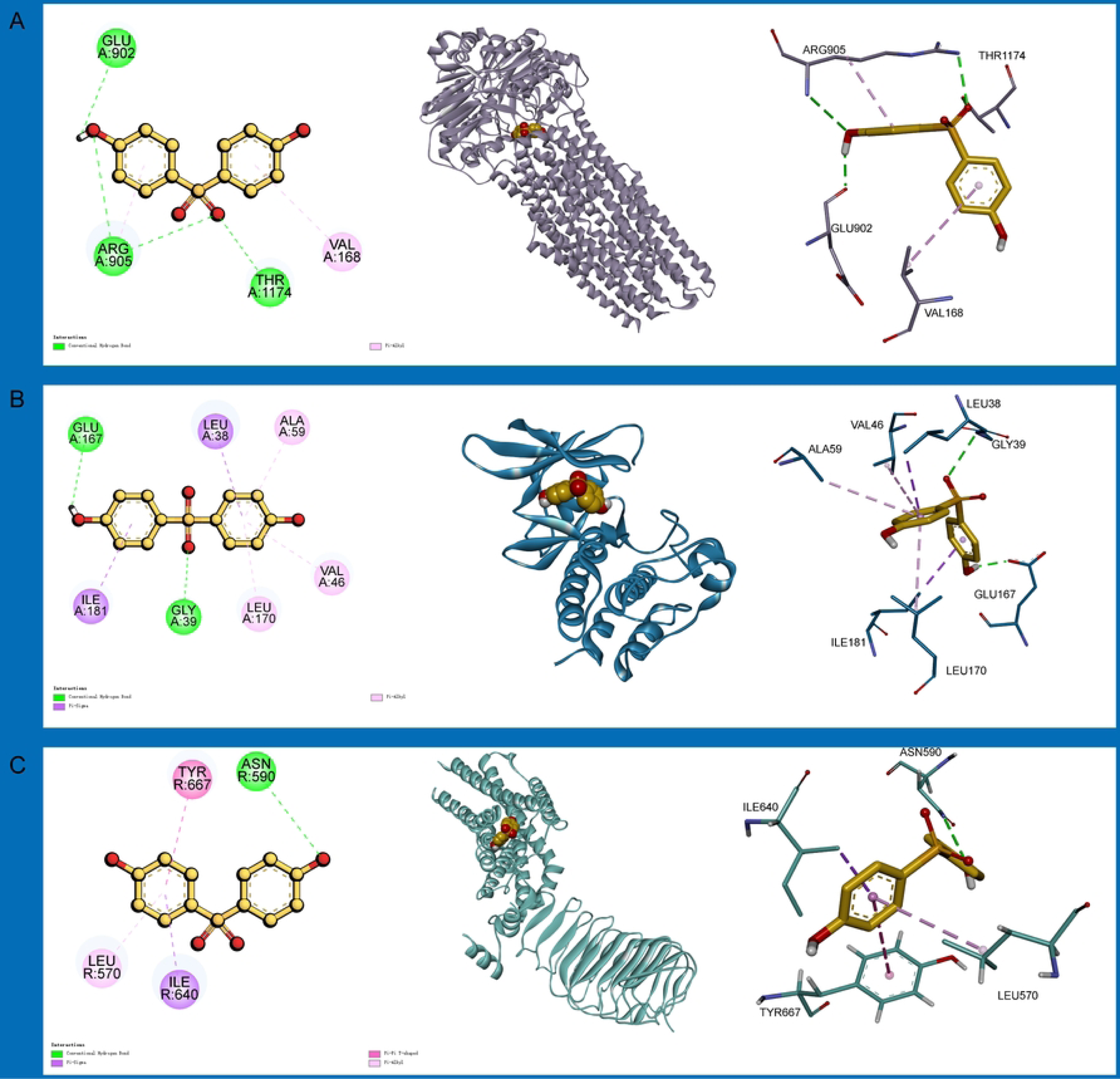

## Notes

### Competing Interest Statement

The authors have declared that no competing interests exist

## References

[1] Tasdogan A, Sullivan R J, Katalinic A, et al. Cutaneous melanoma [J]. Nat Rev Dis Primers, 2025, 11(1): 23.

[2] Long G V, Swetter S M, Menzies A M, et al. Cutaneous melanoma [J]. Lancet, 2023, 402(10400): 485–502.

[3] Sung H, Ferlay J, Siegel R L, et al. Global Cancer Statistics 2020: GLOBOCAN Estimates of Incidence and Mortality Worldwide for 36 Cancers in 185 Countries [J]. CA Cancer J Clin, 2021, 71(3): 209–49.

[4] Gershenwald J E, Scolyer R A, Hess K R, et al. Melanoma staging: Evidence-based changes in the American Joint Committee on Cancer eighth edition cancer staging manual [J]. CA Cancer J Clin, 2017, 67(6): 472–92.

[5] Robert C, Long G V, Brady B, et al. Nivolumab in previously untreated melanoma without BRAF mutation [J]. N Engl J Med, 2015, 372(4): 320–30.

[6] Liao C, Kannan K. A survey of bisphenol A and other bisphenol analogues in foodstuffs from nine cities in China [J]. Food Addit Contam Part A Chem Anal Control Expo Risk Assess, 2014, 31(2): 319–29.

[7] Chen Y, Zhang X, Yang L, et al. The role of cord blood lipidomics in the associations of prenatal exposure to bisphenol A and its analogs with infant growth: A birth cohort study [J]. Ecotoxicol Environ Saf, 2025, 305: 119171.

[8] Zhang Q, Li M, Wang P, et al. Integrated analysis reveals the immunotoxicity mechanism of BPs on human lymphocytes [J]. Chem Biol Interact, 2024, 399: 111148.

[9] Lebachelier De La Riviere M E, Téteau O, Mahé C, et al. Metabolic status is a key factor influencing proteomic changes in ewe granulosa cells induced by chronic BPS exposure [J]. BMC Genomics, 2024, 25(1): 1095.

[10] Zhou X, Wei C, Liu X, et al. Revealing the role of bisphenol A on prostate cancer progression and identifying potential targets: A comprehensive analysis from population cohort to molecular mechanism [J]. Ecotoxicol Environ Saf, 2025, 296: 118209.

[11] Ju G, Zhan X, Chen X, et al. Bisphenol S enhances the cell proliferation ability of prostate cancer cells by regulating the expression of SDS [J]. Toxicol In Vitro, 2024, 98: 105827.

[12] Zhang D, Zhao K, Han T, et al. Bisphenol A promote the cell proliferation and invasion ability of prostate cancer cells via regulating the androgen receptor [J]. Ecotoxicol Environ Saf, 2024, 269: 115818.

[13] Binnewies M, Roberts E W, Kersten K, et al. Understanding the tumor immune microenvironment (TIME) for effective therapy [J]. Nat Med, 2018, 24(5): 541–50.

[14] Soto A M, Sonnenschein C. Environmental causes of cancer: endocrine disruptors as carcinogens [J]. Nat Rev Endocrinol, 2010, 6(7): 363–70.

[15] Smith E A, Belote R L, Cruz N M, et al. Receptor tyrosine kinase inhibition leads to regression of acral melanoma by targeting the tumor microenvironment [J]. J Exp Clin Cancer Res, 2024, 43(1): 317.

[16] Sobczuk P, Cholewiński M, Rutkowski P. Recent advances in tyrosine kinase inhibitors VEGFR 1-3 for the treatment of advanced metastatic melanoma [J]. Expert Opin Pharmacother, 2024, 25(5): 501–10.

[17] Caetano M M M, Moreira G A, Da Silva M R, et al. Impaired expression of serine/arginine protein kinase 2 (SRPK2) affects melanoma progression [J]. Front Genet, 2022, 13: 979735.

[18] Flaherty K T, Puzanov I, Kim K B, et al. Inhibition of mutated, activated BRAF in metastatic melanoma [J]. N Engl J Med, 2010, 363(9): 809–19.

[19] Rout A K, Dehury B, Parida S N, et al. A review on structure-function mechanism and signaling pathway of serine/threonine protein PIM kinases as a therapeutic target [J]. Int J Biol Macromol, 2024, 270(Pt 1): 132030.

[20] Yue L, Xu Y, Lu P. Serine/Threonine Protein Kinase-3 Promotes Oral Squamous Cell Carcinoma by Activating Ras-MAPK Mediated Cell Cycle Progression [J]. Int J Gen Med, 2023, 16: 3115–24.

[21] Ashi A, Awaji A A, Bond J, et al. Threonine and tyrosine kinase (TTK) mRNA and protein expression in breast cancer; prognostic significance in the neoadjuvant setting [J]. Histopathology, 2025, 86(6): 916–32.

[22] Acuña-Pilarte K, Koh M Y. The HIF axes in cancer: angiogenesis, metabolism, and immune-modulation [J]. Trends Biochem Sci, 2025, 50(8): 677–94.

[23] Zhang H, Kong Q, Wang J, et al. Complex roles of cAMP-PKA-CREB signaling in cancer [J]. Exp Hematol Oncol, 2020, 9(1): 32.

[24] Yusupova M, Zhou D, You J, et al. Distinct cAMP Signaling Microdomains Differentially Regulate Melanosomal pH and Pigmentation [J]. J Invest Dermatol, 2023, 143(10): 2019–29.e3.

[25] Latif R, Morshed S A, Ma R, et al. A Gq Biased Small Molecule Active at the TSH Receptor [J]. Front Endocrinol (Lausanne), 2020, 11: 372.

[26] Hawley T S, Riz I, Yang W, et al. Identification of an ABCB1 (P-glycoprotein)-positive carfilzomib-resistant myeloma subpopulation by the pluripotent stem cell fluorescent dye CDy1 [J]. Am J Hematol, 2013, 88(4): 265–72.

[27] Schulz J A, Hartz A M S, Bauer B. ABCB1 and ABCG2 Regulation at the Blood-Brain Barrier: Potential New Targets to Improve Brain Drug Delivery [J]. Pharmacol Rev, 2023, 75(5): 815–53.

[28] Engle K, Kumar G. Cancer multidrug-resistance reversal by ABCB1 inhibition: A recent update [J]. Eur J Med Chem, 2022, 239: 114542.

[29] Yoon S, Kim H S. First-Line Combination Treatment with Low-Dose Bipolar Drugs for ABCB1-Overexpressing Drug-Resistant Cancer Populations [J]. Int J Mol Sci, 2023, 24(9).

[30] Muriithi W, Macharia L W, Heming C P, et al. ABC transporters and the hallmarks of cancer: roles in cancer aggressiveness beyond multidrug resistance [J]. Cancer Biol Med, 2020, 17(2): 253–69.

[31] Chaudhary P M, Mechetner E B, Roninson I B. Expression and activity of the multidrug resistance P-glycoprotein in human peripheral blood lymphocytes [J]. Blood, 1992, 80(11): 2735–9.

[32] Fung K L, Gottesman M M. A synonymous polymorphism in a common MDR1 (ABCB1) haplotype shapes protein function [J]. Biochim Biophys Acta, 2009, 1794(5): 860–71.

[33] Macdonald A, Campbell D G, Toth R, et al. Pim kinases phosphorylate multiple sites on Bad and promote 14-3-3 binding and dissociation from Bcl-XL [J]. BMC Cell Biol, 2006, 7: 1.

[34] Amaravadi R, Thompson C B. The survival kinases Akt and Pim as potential pharmacological targets [J]. J Clin Invest, 2005, 115(10): 2618–24.

[35] Lu J, Zavorotinskaya T, Dai Y, et al. Pim2 is required for maintaining multiple myeloma cell growth through modulating TSC2 phosphorylation [J]. Blood, 2013, 122(9): 1610–20.

[36] Brault L, Gasser C, Bracher F, et al. PIM serine/threonine kinases in the pathogenesis and therapy of hematologic malignancies and solid cancers [J]. Haematologica, 2010, 95(6): 1004–15.

[37] Klee A N, Torchia J A, Freeman G J. Novel Antimurine Thyroid-Stimulating Hormone Receptor Monoclonal Antibodies [J]. Monoclon Antib Immunodiagn Immunother, 2023, 42(3): 109–14.

[38] Slominski A, Wortsman J, Kohn L, et al. Expression of hypothalamic-pituitary-thyroid axis related genes in the human skin [J]. J Invest Dermatol, 2002, 119(6): 1449–55.

[39] Tsoi J, Robert L, Paraiso K, et al. Multi-stage Differentiation Defines Melanoma Subtypes with Differential Vulnerability to Drug-Induced Iron-Dependent Oxidative Stress [J]. Cancer Cell, 2018, 33(5): 890–904.e5.

[40] Hao L, Wu W, Xu Y, et al. LncRNA-MALAT1: A Key Participant in the Occurrence and Development of Cancer [J]. Molecules, 2023, 28(5).

[41] Malakoti F, Targhazeh N, Karimzadeh H, et al. Multiple function of lncRNA MALAT1 in cancer occurrence and progression [J]. Chem Biol Drug Des, 2023, 101(5): 1113–37.

[42] Xu D, Wang W, Wang D, et al. Long noncoding RNA MALAT-1: A versatile regulator in cancer progression, metastasis, immunity, and therapeutic resistance [J]. Noncoding RNA Res, 2024, 9(2): 388–406.

[43] Kalkusova K, Taborska P, Stakheev D, et al. The Role of miR-155 in Antitumor Immunity [J]. Cancers (Basel), 2022, 14(21).

[44] Plummer R, Sodergren M H, Hodgson R, et al. TIMEPOINT, a phase 1 study combining MTL-CEBPA with pembrolizumab, supports the immunomodulatory effect of MTL-CEBPA in solid tumors [J]. Cell Rep Med, 2025, 6(4): 102041.

[45] Li H, Yan L, Li B, et al. Inflammatory factor tumor necrosis factor-α (TNF-α) activates P-glycoprotein (P-gp) by phosphorylating c-Jun and thus promotes transportation in placental cells [J]. Transl Pediatr, 2022, 11(9): 1470–81.

[46] Xue X, Li Z, Zhao J, et al. Advances in the relationship between AP-1 and tumorigenesis, development and therapy resistance [J]. Discov Oncol, 2025, 16(1): 61.

[47] Adesoye T, Tripathy D, Hunt K K, et al. Exploring Novel Frontiers: Leveraging STAT3 Signaling for Advanced Cancer Therapeutics [J]. Cancers (Basel), 2024, 16(3).

